# Differences in neuromusculoskeletal injury and disability rates between US Navy aircraft carrier and amphibious assault ships

**DOI:** 10.1101/2024.06.18.24309118

**Authors:** John J. Fraser, Joshua Halfpap, Michael Rosenthal

**Author notes:** .; Twitter: @NavyPT (John J. Fraser). **Declarations:** Dr Fraser reports grants from Congressionally Directed Medical Research Programs and the Office of Naval Research, outside of the submitted work. In addition, Dr Fraser has a patent pending for an Adaptive and Variable Stiffness Ankle Brace, U.S. Provisional Patent Application No. 63254,474. **Prior Presentation.** None. **Clinical Trial Registration:** Not applicable. **Sources of Support / Funding Sources:** None. **Competing Interests:** None. **Institutional Review Board (Human Subjects):** The study protocol was approved by the Naval Health Research Center Institutional Review Board in compliance with all applicable Federal regulations governing the protection of human subjects. Research data were derived from an approved Naval Health Research Center Institutional Review Board protocol, number NHRC.2022.0201-NHSR. **Institutional Animal Care and Use Committee (IACUC):** Not applicable. **Individual Author Contributions:** JJF was responsible for study conception, methodological development, and data analysis. All authors contributed to the data interpretation, manuscript development, critical revision, and final approval of the study. **Data Availability:** All data relevant to the study are included in the article or are available as supplementary files. **Disclaimer:** The opinions and assertions expressed herein are those of the author(s) and do not reflect the official policy or position of the Uniformed Services University of the Health Sciences or the Department of Defense. **Institutional Clearance:** Institutional clearance approved.

## Abstract

**Introduction:** Musculoskeletal injuries (MSKI) are the most common clinical condition in the military that affect medical readiness. Evaluation of MSKI burden and the effects of these injuries on readiness in large deck Navy ships is warranted.

**Materials and Methods:** A retrospective cohort study assessing population-level MSKI rates, limited duty (LIMDU), and long-term disability episode counts of all Sailors assigned to US Navy Aircraft Carriers (CVNs) and Amphibious Assault Ships (LHA/LHD) from November 2016 to February 2023 were extracted from the Musculoskeletal Naval Epidemiological Surveillance Tool. A negative binomial regression and general additive (gaussian) models evaluated the association of ship platform, deployment status, days underway, and sex on MSKI rates and the proportion of cases that resulted in LIMDU, returned-to-duty following LIMDU, or progressed to long-term disability.

**Results:** Sailors attached to CVNs contributed a mean 17893.8±23280.6 person-months, with those attached to LHA/LHDs contributing an average 5981.8±8432.7 person-months. Aboard CVNs, MSKI occurred at a rate of 0.30±0.16/1000 person-months while deployed and 0.64±0.31/1000 person-months in homeport. Aboard LHA/LHDs, Sailors incurred MSKI at a rate of 0.59±0.58/1000 person-months while on deployment and 1.24±0.68/1000 person-months in homeport. Among Sailors aboard CVNs, LIMDU occurred in 7.95±7.75% of MSKI cases while deployed and 5.13±5.26% while in homeport. Aboard LHA/LHDs, 8.57±13.42% of MSKI cases were placed on LIMDU while deployed and 4.95±5.27% while in homeport. In the multivariable assessment of LIMDU, being deployed underway was a significant factor (B=3.62 p=.03, variance explained=3.86%). Sailors that were female and served aboard LHA/LHDs returned to full duty at a significantly greater frequency compared to their male counterparts and Sailors serving aboard CVNs. None of the independent variables evaluated were associated with long-term disability.

**Conclusion:** The findings in the current study demonstrate the substantial burden of MSKI aboard large deck ships, both in homeport and while deployed. Inclusion of a PT aboard LHA/LHDs, like the CVN, may help to prevent and mitigate the effects of MSKI through early access to specialized care and integral injury prevention and performance optimization methods.

## INTRODUCTION

Musculoskeletal injuries (MSKI) are the most common clinical condition in the military.^1^ These injuries frequently result in limited activity and restricted ability of the servicemember to perform military duties, with diminished capability of the unit to meet mission objectives.^2^ While this is salient while in garrison or in homeport, where servicemembers and their units train in preparation for deployment, this becomes even more relevant when servicemembers deploy. While servicemembers have access to specialized medical care while in garrison or homeport, medical services for MSKI are curtailed while underway and typically limited to primary care in all ship platforms except for aircraft carriers (CVNs). When contextualized with the increased exposure to environmental and manmade hazards in the operational setting, the importance of clinicians with the knowledge, skills, and abilities needed to prevent and mitigate the effects of MSKI cannot be overstated.^3^

Operational environments are an important consideration for MSKI in the US Navy, where Sailors function in the industrial setting of modern large deck warships. Ship construction (e.g. steel decks), negotiating steep ladders, and unique occupational exposures experienced by Sailors assigned to ship’s company are a few factors that may contribute to MSKI. CVNs are a capital warship in the US Navy, providing a mobile aviation strike capability used for power projection in meeting national security objectives. Ship’s company assigned to CVNs consists of more than 3000 Sailors, with a medical capability that includes a Navy Physical Therapist as part of the crew.^4^ This clinician provides not only specialized and timely evaluation and management of MSKI, but also serves as the health promotion officer leading ship and strike group-wide prevention and health optimization programs while in port and during the ∼7-month deployments.^4^ This officer is assigned on two-year orders typically prior to deployment, while underway, and during dwell periods. During extended shipyard maintenance, this billet may be gapped to meet other operational needs as the ship’s company relies upon the nearby branch medical clinic for management of MSKI.

The ship most comparable to CVNs in the US fleet is the amphibious assault ship, consisting of the LHA and LHD classes. This ship’s company consists of approximately 1000 Sailors. The mission of the LHA/LHD, like CVNs, is to forward project Marine aviation and ground strike capabilities ashore. The ship’s medical capability, with the exception of two proof of concept projects conducted since 2010,^3,5^ does not typically include a physical therapist.^6^ Ship’s company must rely on primary care practitioners for the management of MSKI while underway or the local branch medical clinic while in homeport.^6^

In previous studies of MSKI, the effects of Navy Physical Therapists aboard CVNs have been centered on measures of injury frequency derived from patient encounters and estimated cost savings from prevented medical evacuations.^3,7–9^ Based on the limitations of the study designs and the potential for bias, there is a need for empirical evaluation of ship-based physical therapists compared to a reference ship that does not have this medical capability. Therefore, the purpose of this study was to evaluate the effects of ship platform on MSKI rate, limited duty (LIMDU), and long-term disability in Sailors assigned to CVNs and LHA/LHDs. Since there is a potential for sex-related differences in care-seeking for injury^10^ and different environmental factors while deployed, these factors will also be evaluated.

## METHODS

A population-based epidemiological retrospective cohort study of all active-duty Sailors assigned to CVNs and LHA/LHDs from November 2016 to February 2023 was performed to assess how ship platform, deployment status, and sex is associated with MSKI rates. In addition to characterizing the MSKI burden aboard large deck ships, the proportion of MSKI cases placed on limited duty (LIMDU), LIMDU cases returned to duty and those who progressed to long-term disability (disability evaluation system [DES] referral) were evaluated. The Strengthening the Reporting of Observational Studies in Epidemiology (STROBE) Statement was used to guide reporting.^11^

MSKI episodes, LIMDU, and long-term disability counts were extracted from the Musculoskeletal Naval Epidemiological Surveillance Tool (MSK NEST) (Navy Bureau of Medicine & Surgery, Falls Church, VA). This surveillance tool, which has been used in prior study of MSKI in the Navy and Marine Corps,^10^ leverages existing validated databases, such as the Military Health System Data Repository (MDR), LIMDU Sailor and Marine Readiness Tracker System (LIMDU SMART), Disability Evaluation System (DES) and Defense Manpower Data Center Reporting System to report injury burden in a deidentified, aggregated database. MSKI episodes within the MSK NEST were derived from healthcare encounters in the MDR and defined using the Army Public Health Center’s Taxonomy for Musculoskeletal Injuries based on an International Classification of Diagnosis, Tenth Revision (ICD-10) classification.^4^ Individuals with repeat visits for the same diagnosis in a single care episode were only counted once. The database does not include any personal identifiable health information. This study was approved as non–human-subjects research by the Institutional Review Board at the U.S. Naval Health Research Center (NHRC.2022.0201.NHSR).

Since military end strength fluctuates annually due to attrition and recruitment of replacements,^12^ the population at risk was a dynamic cohort. Rate of MSKI was calculated and normalized to the subpopulation at risk (with consideration to ship platform, deployment status, and sex) during the 6-year study epoch. A multivariable negative binomial regression was performed to evaluate the association of ship platform, deployment status, days underway, and sex on MSKI rates. General additive (gaussian) models were used to evaluate ship platform, deployment status, days underway between ports, and sex on the percentage of cases that resulted in LIMDU, returned-to-duty following LIMDU, or long-term disability. Since days underway was not a significant factor for any of these analyses, it was removed from the final parsimonious model. The regression analyses were performed using the ‘PSCL’ (version 1.5.5.1) ‘MGCV’ (version 1.9), and ‘TidyGAM’ (version 0.2) packages on R (version 4.3.1, The R Foundation for Statistical Computing, Vienna, Austria). LHA/LHD served as the reference in the assessment of ship platform. Homeport was selected as the reference for deployment status. Male Sailors served as the reference group in the assessment of sex. The level of significance was *p<*0.05 for all analyses. Statistical significance was evaluated using the convergence CIs that did not cross 1.00 and the *p*-value reported in the regression analysis.

## RESULTS

During the study epoch, CVNs deployed 56 times (mean 167.0±69.1 days) and had 74 dwell periods in homeport (mean 545.8±572.3 days). LHA/LHDs deployed 40 times (mean 148.0±69.7 days) and had 54 dwell times in homeport (mean 490.2±388.5 days). There was no significant interaction observed between platform and deployed time (p=.73). Sailors attached to CVNs contributed a mean 17893.8±23280.6 person-months, with those attached to LHA/LHDs contributing an average 5981.8±8432.7 person-months.

### MSKI Rate

Among Sailors aboard CVNs, MSKI occurred at a rate of 0.30±0.16/1000 person-months while deployed and 0.64±0.31/1000 person-months in homeport (**Figure 1**). Aboard LHA/LHDs, Sailors incurred MSKI at a rate of 0.59±0.58/1000 person-months while on deployment and 1.24±0.68/1000 person-months in homeport. In the multivariable assessment of MSKI rate, service aboard aircraft carriers (p<.001), being deployed (p<.001), and male sex (p<.001) were found to have significant and large magnitude protective effects (RR range 0.51-0.77) (**Figure 1**).

**Figure 1.**
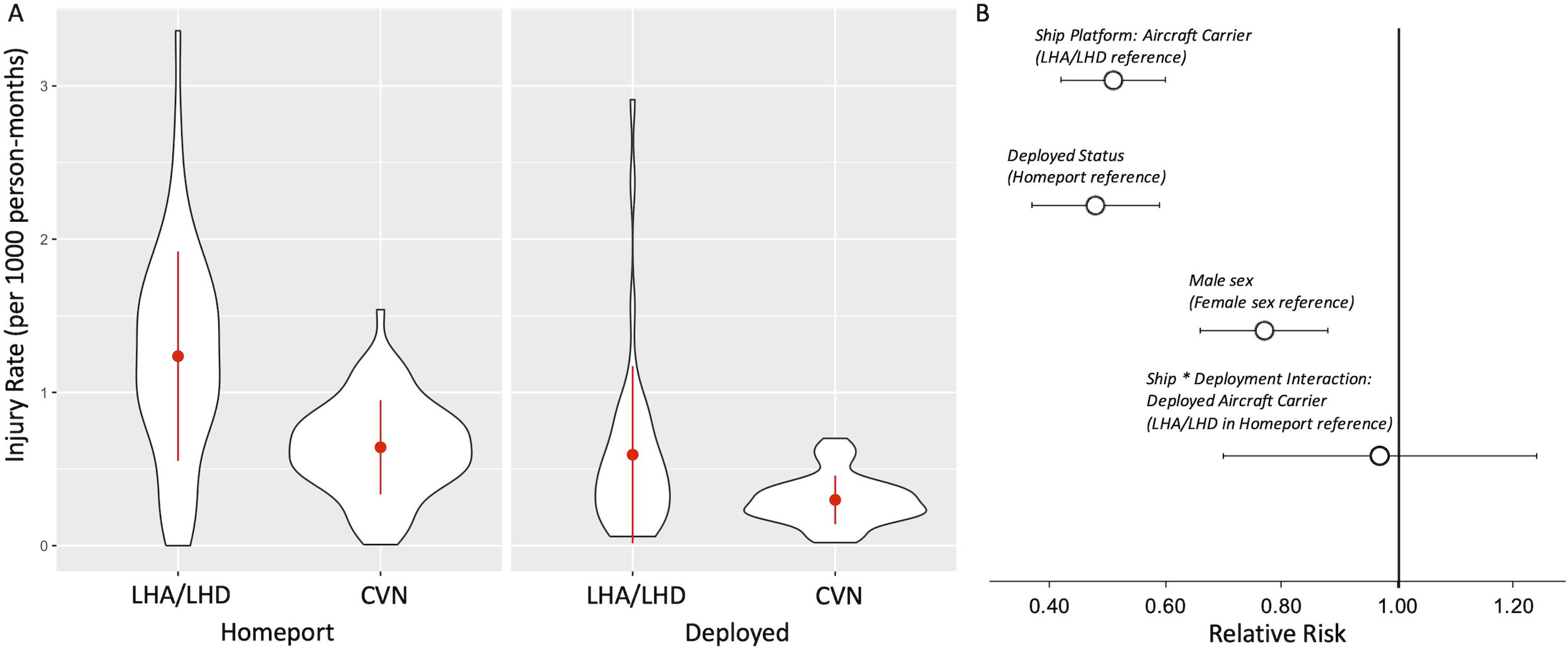
(a) Descriptive statistics and (b) results of the negative binomial regression evaluating ship platform, deployment status, sex, and ship by deployment interaction on musculoskeletal injury rates aboard US Navy amphibious assault ships (LHA/LHD class) and aircraft carriers (CVN class) from November 2016 to February 2023.

### MSKI-related LIMDU and Long-term Disability

Among Sailors aboard CVNs, LIMDU occurred in 7.95±7.75% of MSKI cases while deployed and 5.13±5.26% while in homeport (**Figure 2**). Aboard LHA/LHDs, 8.57±13.42% of MSKI cases were placed on LIMDU while deployed and 4.95±5.27% while in homeport. In the multivariable assessment of LIMDU, being deployed underway was a significant factor (B=3.62 p=.03, variance explained=3.86%) (**Figure 2**). In the assessment of Sailors returned to full duty following LIMDU (CVN: deployed=61.33±43.29%, homeport=40.79±38.76%; LHA/LHD: deployed=78.58±37.81%, homeport=40.60±42.27%), service aboard LHA/LHD (B=12.42, p=.02), being deployed (B=24.07, p<.001), and female sex (B=14.92 p=.006) had greater proportion of cases returned to full function (variance explained=13.00%). Of the Sailors who progressed to long-term disability (DES referral) (CVN: deployed=1.15±2.85%, homeport=0.75±3.00%; LHA/LHD: deployed=0.48±1.72%, homeport=0.25±0.55%), none of the study predictors were significant. There were no other significant findings observed.

**Figure 2.**
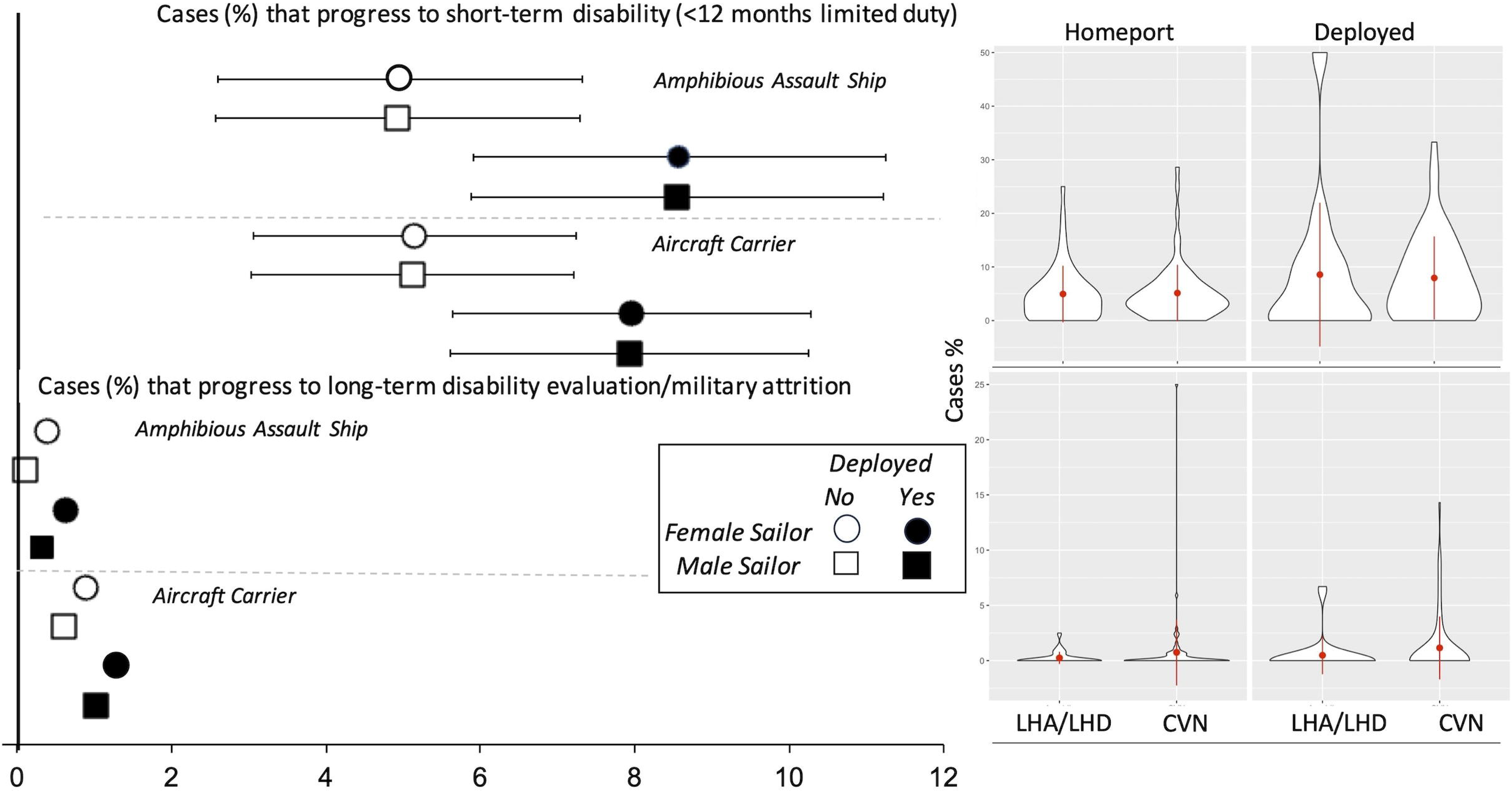
Point estimates and 95% confidence intervals detailing the mean proportion of Sailors on musculoskeletal injury-related short and long-term disability aboard US Navy amphibious assault ships (LHA/LHD class) and aircraft carriers (CVN class), stratified by sex, platform, and deployment status, from November 2016 to February 2023.

**Figure 3.**
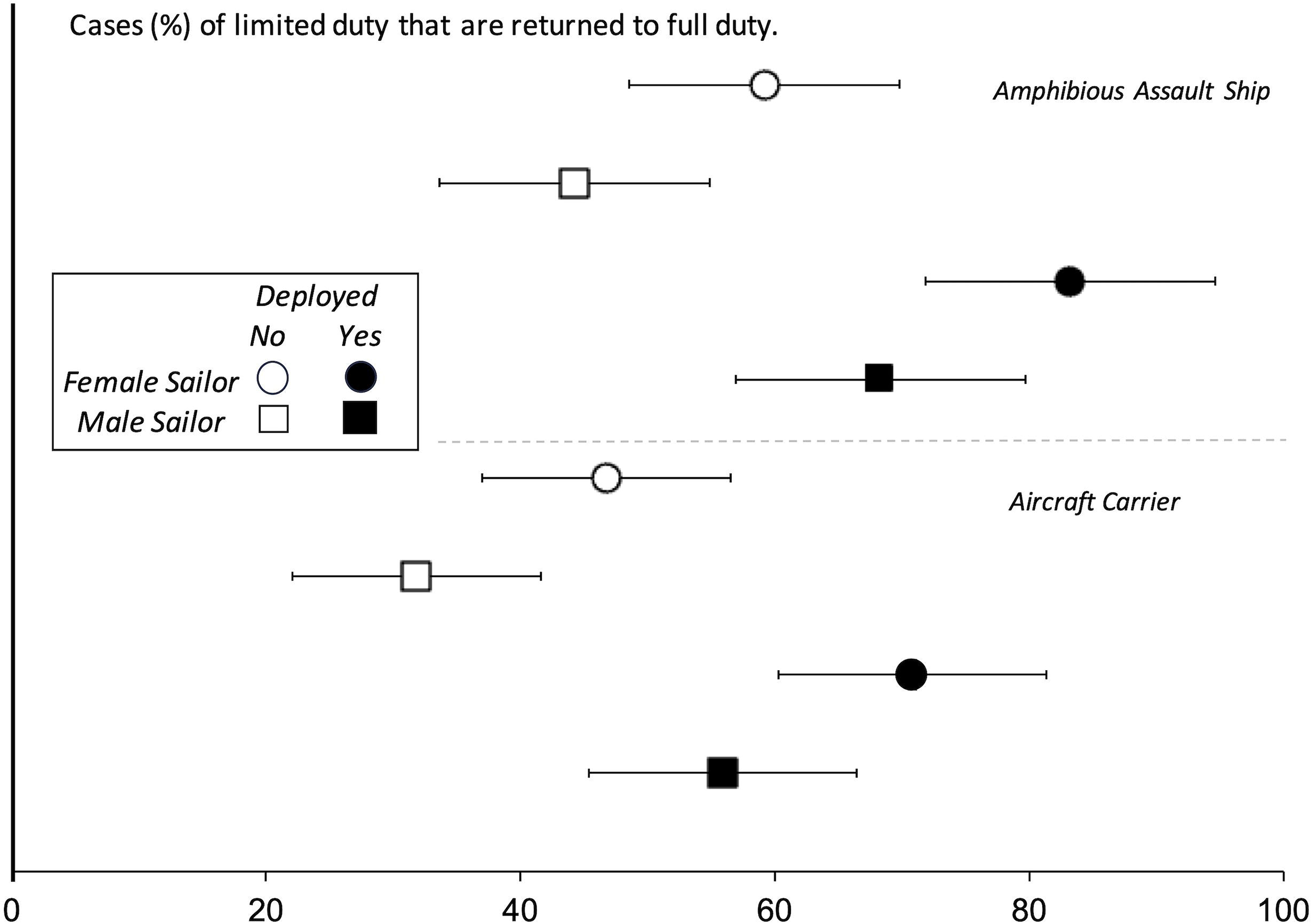
Point estimates and 95% confidence intervals detailing the mean proportion of Sailors returned to full duty following musculoskeletal injury-related limited duty (short-term disability) aboard US Navy amphibious assault ships (LHA/LHD class) and aircraft carriers (CVN class), stratified by sex, platform, and deployment status, from November 2016 to February 2023.

## DISCUSSION

The primary findings of this study were that service aboard an LHA/LHD, being in homeport, and female sex were salient risk factors for MSKI. Being deployed was significantly associated with Sailors being placed on LIMDU and returned to duty following a LIMDU period. Sailors that were female and served aboard LHA/LHDs returned to full duty at a significantly greater proportion compared to their male counterparts and Sailors serving aboard CVNs. None of the independent variables evaluated were associated with long-term disability.

### MSKI Rate

Sailors assigned to CVNs were 0.51 times less likely to have a MSKI compared to LHA/LHDs. Key functional and structural differences between the ships include the number and type of personnel assigned to the ship, the size of the ship, and capabilities/mission requirements. These differences may result in different types of occupational exposure and hazards. For example, CVNs have a unique catapult system used to launch aircraft and a nuclear reactor, with LHA/LHDs having a well deck capable of carrying, flooding, and launching various amphibious assault craft. Sociocultural differences between these types of ships also likely exist which may influence differences in MSKI. Another difference is U.S. Navy Physical Therapists have been permanently assigned to CVNs dating back to 2002.^9^ The directives that fully detail shipboard medical manning and procedures can be found in the referenced Navy policy.^4,6^ Physical Therapists provide the primary evaluation, management, and treatment of MSKI onboard CVNs. Additionally, the ship’s physical therapists are responsible to lead comprehensive health promotion programs as part of their duties.^4^ It is plausible that MSKI prevention and performance programs administered by the ship’s physical therapist may have contributed to the protective effect observed aboard CVNs.

Females Sailors demonstrated a significantly higher rate of MSKI compared to their male counterparts. This finding is consistent with a prior study of sex as a factor for MSKI in Sailors using the MSK NEST database.^10^ Observed sex-related differences in MSKI risk can be attributed to intrinsic biomechanical, physiological, and psychological factors, as well as extrinsic social and environmental factors that include service culture, occupational exposure, and rank.^10,13^ Social determinants such as care-seeking behaviors, stigma, the pressure to work harder than their male counterparts, and other social pressures that may be sex related, a supposition that warrants further investigation. ^3,10,13^

### LIMDU and Long-term Disability

The Navy Medicine LIMDU program is the primary method for managing the medical care for ill and injured active Sailors to ensure a medically ready force.^14^ In the shipboard and deployed environment, medical readiness is more heavily affected when Sailors are placed in a LIMDU status. If a Sailor is unable to perform activities of daily living or occupational-related tasks, the LIMDU policy necessitates the transfer of the Sailor off the ship. This may explain why LIMDU placement rates were marginally (∼3-4%) higher while deployed compared to when the ship was in homeport, regardless of ship type. While we did not characterize the type or severity of MSKI that warranted LIMDU placement in the current study, it is highly plausible that these factors were salient in the determination of LIMDU placement. Given the hazards of the industrial environment of a warship and operations at sea, the regulations that govern the suitability for a Sailor to function aboard ship will take precedence, regardless of ship type and the specialists aboard.^4,6^

### Return to Full Duty following LIMDU

When controlling for deployment status, days underway, and sex, a significantly higher proportion of Sailors assigned to a deployed LHA/LHDs returned to full duty following LIMDU (mean difference ∼17%) compared to those aboard deployed CVNs. The mean proportion of Sailors that returned to full duty while in homeport were similar in both ship platforms (∼41%). The greater return to duty in deployed Sailors may be due to motivations, career advancement, or financial incentives associated with being deployed. Deployment, as part of a full scope military operation, is the culmination of service and the numerous hours of labor and training needed during preparation for operations at sea. It is plausible that for many, minimization of persistent pain and disability and prioritization of the needs of the mission over self may explain this finding.^3,13^

We observed a higher rate of return to duty in female Sailors. This may be in part due to greater healthcare utilization while deployed. In a prior study of healthcare use during deployment, female Sailors were found to use ship’s medical services at a rate 9.2 times higher than their male counterparts.^15^ It is plausible that female Sailors with greater healthcare utilization more readily benefitted from the guidance, relation-inferred efficacy, and social support provided by the ship’s medical staff during recovery.^16^ Another explanation for this finding may be a function of selection of military occupation and the associated physical demands.^13^ Female service members historically have gravitated toward military occupations with fewer physical demands, with administration, healthcare, and supply being the most common fields.^17^ It is possible that women serving in fields with fewer physical requirements can more readily return to full duty, even if there is persistent pain and impairment following MSKI. These suppositions warrant future investigation.

### Policy and Practice Implications

The findings in the current study demonstrate the substantial burden of MSKI aboard large deck ships, both in homeport and while deployed. Inclusion of a PT aboard LHA/LHDs, like the CVN, may help to prevent and mitigate the effects of MSKI through early access to specialized care and integral injury prevention and performance optimization methods. This concept was evaluated in two proof of concept projects, the first which was conducted in 2010.^3,5^ The Sports Medicine on the Battlefield model promotes early access to physical therapist care and has found to be effective in expediting return to duty and optimizing recovery during theatre medical operations.^18^ More recently, this model has been employed and demonstrated to be effective in the Military Health System.^19^ In the current era of declining eligibility for military service, failure to meet annual recruiting targets, and fluctuating retention, efforts to fortify Naval capital warships with resources that support the adage “humans are more important than hardware” warrants recurring visitation.^3,13^

### Limitations

There are limitations to this study. First, this study relied on deidentified, aggregated data provided in the NEST database. While this allowed us to ascertain population-level burden of MSKI rate, short-term LIMDU, and long-term disability, the lack of individual-level data precluded the assessment of comorbidities and other intrinsic factors that may have contributed to the outcomes under study. This study relied on health encounters in the electronic medical record to ascertain MSKI. Sailors who did not sought care or received undocumented care following MSKI were not captured in this evaluation, so the reported burden is likely underestimated in the current study. In the evaluation of ship platform, we used the ship class LHA/LHD as the most comparable reference to that of the CVN in the fleet (based on size and mission set) to evaluate the effectiveness of the physical therapist aboard ship. While there are inherent limitations for this choice (with the amphibious ships having slightly different mission capabilities and requirements that likely influences exposure), this is the only contrast to our knowledge available for this assessment. Future studies evaluating differences in functional capacity requirements, occupational exposures, and environmental hazards across platforms would help to better elucidate if this influenced the findings in the current study. Lastly, we could not qualify the type and severity of injury incurred by the Sailors, nor the clinician type making the determination for LIMDU and return to duty. Given that the responsibility for the medical needs of Sailors aboard ship is with the Senior Medical Officer (typically a board-certified family, aviation health, or occupational medicine trained physician),^4,6^ retention of a Sailor aboard board ship is often a complex decision that balances the needs of the Sailor, the needs of the ship/mission, and contextualized by the social and environmental factors when making risk decisions. While the physical therapist aboard CVNs have a role in providing expert counsel to the Senior Medical Officer pertaining to disposition, it was not possible to evaluate these potential contextualizing factors. While Sailors with MSKI returned to duty at approximately the same frequency in both ship platforms, differences in the long-term health-related quality of life and function of these Sailors could not be ascertained in this study.

## CONCLUSION

MSKI was common in Sailors assigned to large deck ships, with service aboard an LHA/LHD, being in homeport, and female sex identified as salient risk factors. Being deployed was significantly associated with Sailors being placed on LIMDU and returned to duty following a LIMDU period. Sailors that were female and served aboard LHA/LHDs returned to full duty at a significantly greater proportion compared to their male counterparts and Sailors serving aboard CVNs. None of the independent variables evaluated were associated with long-term disability.

These findings can be used to better plan prevention efforts and medical care needed aboard ship following MSKI. The findings in the current study demonstrate the substantial burden of MSKI aboard large deck ships, both in homeport and while deployed. Inclusion of a PT aboard LHA/LHDs, like the CVN, may help to prevent and mitigate the effects of MSKI through early access to specialized care and integral injury prevention and performance optimization methods.

### Patient and Public Involvement

The results of this study were provided to the US Navy Bureau of Medicine and Surgery Neuromusculoskeletal Clinical Community for dissemination and translation.

## Data Availability

All data relevant to the study are included in the article or are available as supplementary files.

## Acknowledgements

None

## Notes

### Funding Statement

This study did not receive any funding.

### Author Declarations

The study protocol was approved by the Naval Health Research Center Institutional Review Board in compliance with all applicable Federal regulations governing the protection of human subjects. Research data were derived from an approved Naval Health Research Center Institutional Review Board protocol, number NHRC.2022.0201-NHSR.

